# Occupation Recognition and Exploitation in Rheumatology Clinical Notes: Employing Deep Learning Models for Named Entity Recognition and Knowledge Discovery in Electronic Health Records

**DOI:** 10.1101/2024.05.08.24306389

**Authors:** Alfredo Madrid-García, Inés Pérez-Sancristóbal, Leticia-Leon, Lydia-Abásolo, Benjamín Fernández-Gutiérrez, Luis Rodríguez-Rodríguez

**Affiliations:** Grupo de Patología Musculoesquelética. Hospital Clínico San Carlos. Instituto de Investigación Sanitaria San Carlos (IdISSC), Prof. Martin Lagos s/n, Madrid, 28040, Spain; Facultad de Medicina, Universidad Complutense de Madrid, Pl. de Ramón y Cajal, s/n, Moncloa - Aravaca, Madrid, 28040, Spain

**Keywords:** artificial intelligence, transformers, named entity recognition, real-world data, rheumatology, natural language processing, occupations, electronic health record, work disability, occupational medicine

## Abstract

Occupation is considered a Social Determinant of Health (SDOH) and its effects have been studied at multiple levels. Although the inclusion of such data in the Electronic Health Record (EHR) is vital for the provision of clinical care, specially in rheumatology where work disability prevention is essential, occupation information is often either not routinely documented or captured in an unstructured manner within conventional EHR systems. Encouraged by recent advances in natural language processing and deep learning models, we propose the use of novel architectures (i.e., transformers) to detect occupation mentions in rheumatology clinical notes of a tertiary hospital, and to whom those occupations belongs. We also aimed to evaluate the clinical and demographic characteristics that influence the collection of this SDOH; and the association between occupation and patients’ diagnosis. Bivariate and multivariate logistic regression analysis were conducted for this purpose.

A Spanish pre-trained language model, RoBERTa, fine-tuned with biomedical texts was used to detect occupations. The best model achieved a F1-score of 0.725 identifying occupation mentions. Moreover, highly disabling mechanical pathology diagnoses (i.e., back pain, muscle disorders) were associated with a higher probability of occupation collection. Ultimately, we determined the professions most closely associated with more than ten categories of muscu-loskeletal disorders.

**Highlights:**

- Deep learning models hold significant potential for structuring and leveraging information in rheumatology
- Diagnoses related to highly disabling mechanical pathology were associated with a higher probability of occupation collection
- Cleaners, helpers, and social workers occupations are linked to mechanical pathologies such as back pain

## 1. Introduction

The relationship between occupation and health has been studied for more than a century [1]. Nowadays, the benefits of incorporating occupation-related information in the Electronic Health Record (EHR), such as a more accurate diagnosis or better health outcomes, have been widely characterized [2, 3], and promoted by agencies such as the National Institute for Occupational Safety and Health (NIOSH) or European Agency for Safety and Health at Work (EU-OSHA) [4]. Although the relevance of this Social Determinant of Health (SDOH) [5] has been identified and studied at multiple health levels (i.e., mental health [6], health inequality [7], or self-rated health [8]), its study is overshadowed by other SDOH including gender, race, or ethnicity [9, 10]; compromising its content [11], variability [12] and quality [13]. Moreover, most SDOH are stored as free-text unstructured data [14] making them difficult to handle and use.

In the field of Rheumatic and Musculoskeletal Diseases (RMDs), the need to accurately capture occupation information is crucial for the provision of care and to promote prevention and intervention activities that can reduce work disability and sick leave [15]. For instance, recent studies have shown an association between occupation and different inflammatory and non-inflammatory musculoskeletal diseases, such as low back pain [16], Osteoarthritis of the knee (OAk) [17], epicondylitis [18], fibromyalgia severity [19], systemic lupus erythematosus [20], Rheumatoid Arthritis (RA) [21, 22], and presence of anti-citrullinated protein antibodies (ACPA) [23] and anti-neutrophil cytoplasmic antibodies (ANCA) [24].

Automatic detection of information in free-text notes using Natural Language Processing (NLP) techniques and its classification in pre-defined categories is called Named Entity Recognition (NER). This information extraction task has benefited from recent advances in Deep Learning (DL) architectures such as transformers. Briefly, these models are trained on a vast amount of data in an unsupervised way to learn the general structure of a language and its vocabulary [25, 26]. Once trained, the weights of the neurons comprising the model are updated using task-specific data in a process called fine-tuning. In this way, models learn to solve specific tasks in a particular domain. Transformers have been applied in the past in rheumatology for the classification of temporal artery biopsy reports [27], or to identify and classify medical entities in clinical notes related to RA [28]. Until today, studies that focus on the patient’s occupation are scarce, and even more, studies that attempt to identify occupation mentions in EHR using advanced DL approaches, for knowledge discovery. To address this gap and with the aim of characterizing the patient’s occupation, we proposed the use of transformers for occupation NER in a rheumatology departmental EHR. By structuring occupational data, we pursue to characterised the relevance of this SDOH in rheumatology. Therefore, the objective of this study is three-fold:

1. To assess the performance of NER models when extracting occupation mentions in rheumatology clinical notes.
2. To describe the demographic and clinical characteristics that influence the collection of occupation-related information.
3. To analyze the association between occupation and patient’s diagnosis.

## 2. Material and Methods

### 2.1. Hospital Clínico San Carlos Musculoskeletal Cohort and inclusion criteria

Clinical narratives from the Hospital Clínico San Carlos Musculoskeletal Cohort (HCSC-MSKC) were retrieved. The HCSC-MSKC is a routine clinical practice cohort that includes more than 117,000 visits and 35,000 subjects seen at the rheumatology outpatient clinic from 1^st^ April 2007 to 30^th^ November 2017. More details about this data source can be found elsewhere [29]. The sex-age distribution of patients belonging to this cohort at their first visit is shown in Supplementary Figure 1.

Only visits containing free-text information from patients with free-text information in the first visit were retrieved. After a preliminary analysis and to avoid selection bias, only patients classified as *active* in the *labour situation* structured variable from the EHR and aged between 16 and 65 were included.

### 2.2. Methodology

#### 2.2.1. Methodology for performance of NER models objective

In a previous study [25], we developed a model for occupation detection, understood as a sequence-labelling NER problem; and another model for the attribution of such occupation to the different stakeholders involved in the healthcare process. Both of them are accessible through Hugging Face [30]. They are RoBERTa-based [31] models, trained with Spanish biomedical texts and fine-tuned using data from MEDDOPROF corpus [32]. MEDDOPROF is a public corpus consisting of 1,844 Spanish clinical case reports with annotations for occupations (i.e., occupations that provide a person with an income or livelihood), working status, and activities (i.e., non-remunerated professions); as well as annotations for to whom the occupation belongs, namely, patient, family member, health professional, or others. Hence, the corpus contained a total of seven classes. More details about this corpus and the model development are shown in Supplementary Material *MEDDOPROF corpus description* and *Model development* sections.

In the present study, 2,000 first visit clinical notes from the HCSC-MSKC were randomly chosen for annotation to build a gold standard, and to evaluate the performance of the previous models in a real-world scenario. These 2,000 notes were annotated (i.e., the occupations and the stakeholders contained in the EHR are manually highlighted using software tools) by two annotators, AMG and IPS, using brat rapid annotation tool (BRAT) and following MEDDOPROF corpus annotation guidelines [33]. No data from the HCSC-MSKC was used for fine-tuning or training the models. The Inter-Annotator Agreement (IAA) F1-score was measured using the *bratiaa* python package, accessible through GitHub [34]. Discrepancies between annotators were resolved by consensus.

Once the gold standard set was built, inference was made locally. The evaluation metrics used to assess the performance of the models were precision, recall and F1-score [35], microaveraged. The overall values of these metrics were calculated using the MEDDOPROF task evaluation script, also accesible through GitHub [36]. The values for each entity were computed using the sequence labeling evaluation (seqeval) Python library [37]. Confusion matrices at the token level are provided in the Supplementary Material, *Supplementary tables* section.

After analysing the models’ performance, predictions were made on the rest of the HCSC-MSKC notes. Overlapping entities (i.e., same text recognised by the two models), classified as “profession-patient”; as well as, entities classified as “profession” (i.e., the text is only recognised by the first model) or “patient” were retrieved, as we found that we could recover approximately 10% of professions attributable to patients, see Supplementary Material *Annotation process and gold standard* and *Predictions in the HCSC-MSKC notes* sections.

#### 2.2.2. Methodology for demographic and clinical characteristics that influence occupation collection objective

Once the predictions were made and manually reviewed, the European Skills, Competencies, Qualifications and Occupations (ESCO) classification version 1.1.1 was used to normalize the patients’ occupations. Visits with more than one occupation (e.g., past and present occupation or moonlighter) were also considered. Only the first visit where the patient’s occupation was mentioned was obtained, then the nearest neighbor matching method (1:1) was used to identify visits of patients without any occupation mention, with *MatchIt* R package [38], Supplementary Material *Matching visits*. Age, sex, visit number, time since first visit to the rheumatology clinic until current visit and calendar year variables were used to conduct the matching. Logistic regression bivariate and multivariate analyses were conducted to identify clinical and demographic predictors associated with the occupation collection (i.e., occupation collection as the dependent variable, with clinical and demographic factors as independent variables). Firstly, only predictors with a prevalence > 5% in the group of visits with occupation mentions were evaluated, otherwise, they were reclassified into broader categories until the minimum prevalence was reached, see *Supplementary Excel File Predictors Classification*. The predictors used in the matching were not included in the analyses. Secondly, predictors with a p-value < 0.15 in the bivariate analyses were included in the multivariate analyse. On the other hand, the predictors selection in the multivariate analysis was conducted following an hybrid stepwise approach, optimising the Akaike Information Criterion (AIC) value.

#### 2.2.3. Methodology for association between occupation and patient’s diagnosis objective

To assess the relation between professions and diagnoses, we conducted bivariate and multivariate logistic regression analyses, using each disease as the dependent variable and the occupations as independent variables. To this end, ESCO codes were re-classified into categories to ensure a minimum prevalence of 20 mentions per occupation category, see *Supplementary Excel File ESCO*. Only patients visits with occupation mentions were studied. As in the previous objective, predictors (i.e., occupations) with a p-value < 0.15 in the bivariate analyses were included in the multivariate analysis. A multivariate analysis, adjusting by age and sex; and following an hybrid stepwise approach optimising the AIC was performed for each of the 13 disease groups.

Python 3.8.16 was utilized to fine-tune the NER models, and R 4.3.1 was employed in the desriptive and statistical analyses.

### 2.3. Statistical analysis

Structured demographic and clinical variables, including sex, age, visit type, disease, or Quality of Life (QoL) were used to characterise patients and visits with and without occupation mentions. Dichotomous and categorical variables were summarised using proportions. Continuous variables were summarised using the median and first and third quartiles (Q1–Q3).

### 2.4. Ethics board approval and reporting guidelines

HCSC Ethics Review Board approval (23/340-E) was obtained as a retrospective study and waiver of informed consent was obtained for the use of unidentified clinical records. Furthermore, the study was conducted in accordance with the Declaration of Helsinki. MI-CLAIM checklist was used to report the results of this study [39], see *Supplementary Material MI-CLAIM checklist*.

## 3. Results

### 3.1. Performance of NER models in HCSC-MSKC gold standard set

The precision, recall, and the microaveraged F1-score values in the gold standard set, for the occupation detection task, were 0.806, 0.633, and 0.709; for the actors associated with the occupation 0.783, 0.702, and 0.740; and combined 0.795, 0.668, and 0.725. In the gold standard set, only 8.05% of notes (i.e., 161 out of 2,000) contained one “profession-patient” mention and six notes contained more than one “profession-patient” mention. The total number of “profession-patient” mentions was 167, see Supplementary Table 7.

### 3.2. Demographic and clinical characteristics that influence occupation collection

#### 3.2.1. Predictions in the HCSC-MSKC notes

Predictions were made on a total of 117,068 free-text notes from 35,470 patients. From 5,917 visits in which at least one entity was recognised as “patient”, “profession” or “profession-patient”, 3,978 visits (67.22%) from 3,723 patients (10.5%) have a real occupation registered, after conducting a manual review. Hence, only the 3.40% out of the whole HCSC-MSKC notes set have occupation-related information. Supplementary Figure 2 shows the percentage of occupation collection stratified by physician.

Considering the demographics and clinical characteristics of the visits, Table 1, and that only 218 out of the 3,723 (5.86%) patients have more than one visit with an occupation mention, we opted to only analysed the visits in which the occupation was collected for the first time, irrespective of whether it was the patient’s first visit or not. Finally, the rest of the inclusion criteria were applied; see Figure 1.

**Table 1.** Clinical and sociodemographic characteristics of all visits with occupation mentions.

| Characteristics | Visits with<br>occupation mentions (n = 3,978) |
| --- | --- |
| **Women** | 2,619 (65.84%) |
| **Age** | 47.67 (38.98, 54.52) |
| **Visit number** |  |
| 1 | 3,167 (79.61%) |
| 2 | 347 (8.72%) |
| 3 | 125 (3.14%) |
| 4 | 69 (1.73%) |
| >4 | 270 (6.79%) |
| **Time from first visit (days)** | 570.00 (162.00, 1,155.00) |
| **Calendar year** |  |
| 2007 | 99 (2.49%) |
| 2008 | 161 (4.05%) |
| 2009 | 253 (6.36%) |
| 2010 | 317 (7.97%) |
| 2011 | 379 (9.53%) |
| 2012 | 415 (10.43%) |
| 2013 | 489 (12.29%) |
| 2014 | 449 (11.29%) |
| 2015 | 452 (11.36%) |
| 2016 | 509 (12.80%) |
| 2017 | 455 (11.44%) |
| **Labour situation** |  |
| Active | 3,808 (95.73%) |
| Homemaker | 30 (0.75%) |
| Retired | 128 (3.22%) |
| Student | 12 (0.30%) |
| **Distress** |  |
| No distress | 869 (21.85%) |
| Mild | 2,347 (59.00%) |
| Moderate | 741 (18.63%) |
| Severe | 21 (0.53%) |
| **Disability** |  |
| 1 | 1,886 (47.41%) |
| 2 | 1,305 (32.81%) |
| 3 | 490 (12.32%) |
| 4 | 243 (6.11%) |
| 5 | 50 (1.26%) |
| 6 | 4 (0.10%) |
| 7 | 0 (0.00%) |
| **Rosser** | 0.99 (0.98, 0.99) |
| **Training doctor visit** | 185 (4.65%) |

**Figure 1:**
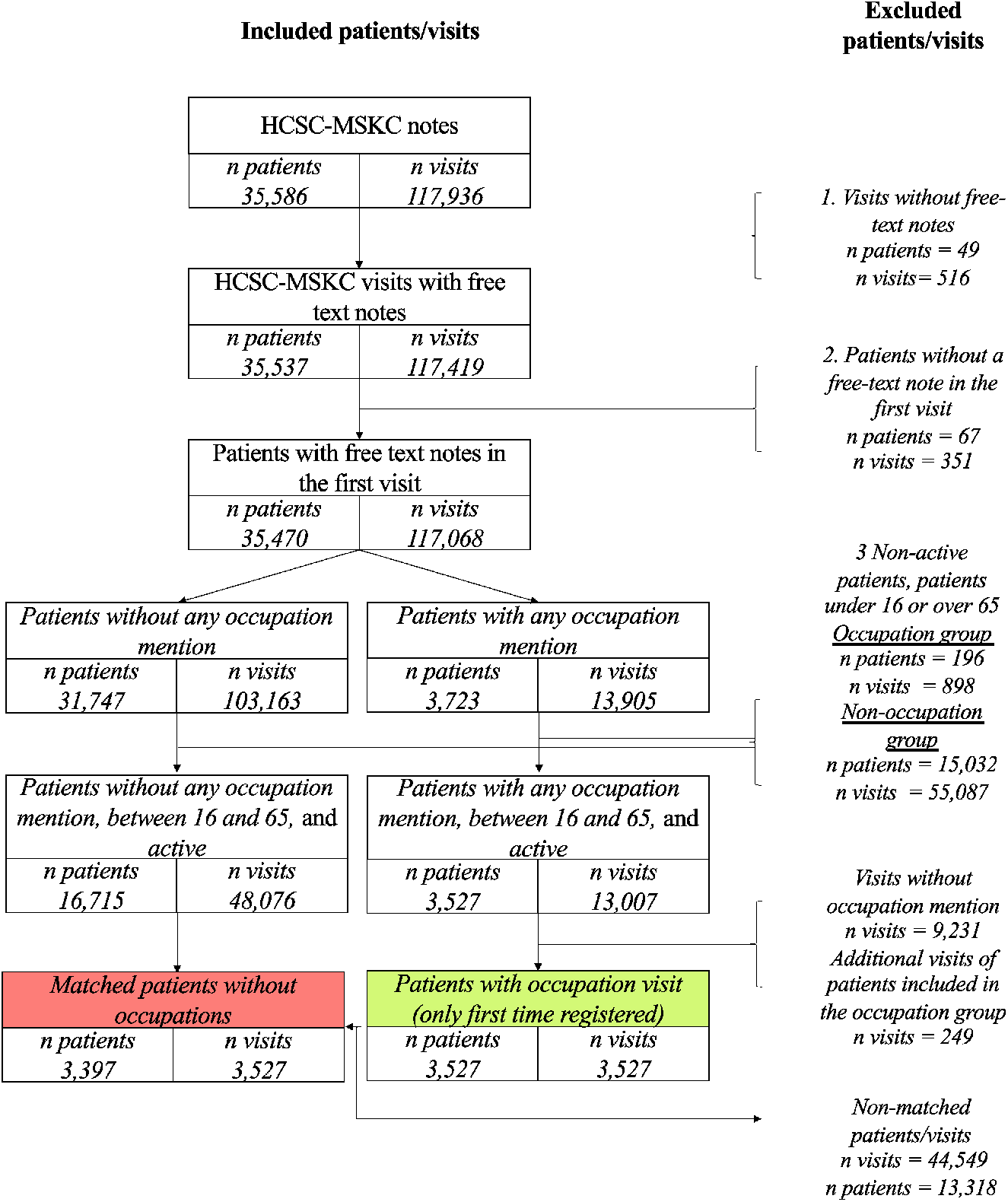
Inclusion criteria diagram

3,527 patients out of 3,723 had at least one occupation mention that met the inclusion criteria. Of these, 515 patients with no occupation information at their first visit had that information collected in a subsequent visit. Supplementary Figure 3 shows a histogram of the time elapsed, days, until occupation collection for these 515 patients.

The 3,527 visits with occupation mentions were compared to 3,527 paired visits of patients who did not have any occupation mention visits, after matching. Table 2 shows the differences between both groups.

**Table 2.** Clinical and sociodemographic characteristics for both groups. ^∤^ excluding first visits.

|  | Matched visits<br>w/o occupation mentions<br>(N = 3,527) | Visits with<br>occupation mentions<br>(N = 3,527) |
| --- | --- | --- |
| **Women** | 2,364 (67.03%) | 2,331 (66.09%) |
| **Age** | 46.78 (37.60, 53.90) | 46.95 (38.36, 53.36) |
| **Visit number** |  |  |
| 1 | 2,913 (82.59%) | 3,012 (85.40%) |
| 2 | 338 (9.58%) | 236 (6.69%) |
| 3 | 83 (2.35%) | 79 (2.24%) |
| 4 | 50 (1.42%) | 45 (1.28%) |
| >4 | 143 (4.05%) | 155 (4.39%) |
| **Time from first visit<sup>†</sup> (days)** | 251.00 (44.00, 933.75) | 581.00 (155.50, 1,140.50) |
| **Calendar year** |  |  |
| 2007 | 109 (3.09%) | 95 (2.69%) |
| 2008 | 196 (5.56%) | 153 (4.34%) |
| 2009 | 166 (4.71%) | 235 (6.66%) |
| 2010 | 254 (7.20%) | 285 (8.08%) |
| 2011 | 334 (9.47%) | 328 (9.30%) |
| 2012 | 379 (10.75%) | 372 (10.55%) |
| 2013 | 444 (12.59%) | 425 (12.05%) |
| 2014 | 420 (11.91%) | 399 (11.31%) |
| 2015 | 404 (11.45%) | 400 (11.34%) |
| 2016 | 441 (12.50%) | 445 (12.62%) |
| 2017 | 380 (10.77%) | 390 (11.06%) |
| **Labour situation** |  |  |
| Active | 3,527 (100.00%) | 3,527 (100.00%) |
| **Distress** |  |  |
| No distress | 687 (19.48%) | 791 (22.43%) |
| Mild | 2,275 (64.50%) | 2,072 (58.75%) |
| Moderate and severe | 565 (16.02%) | 664 (18.83%) |
| **Disability** |  |  |
| No disability | 1,593 (45.17%) | 1,707 (48.40%) |
| Slight disability | 1,344 (38.11%) | 1,121 (31.78%) |
| Disability | 590 (16.73%) | 699 (19.82%) |
| **Rosser** | 0.99 (0.99, 0.99) | 0.99 (0.98, 0.99) |
| **Training doctor visit** | 192 (5.44%) | 148 (4.20%) |
| **Disease** |  |  |
| Back pain | 469 (13.30%) | 842 (23.87%) |
| Tendinitis (upper extremities) | 485 (13.75%) | 711 (20.16%) |
| Pain in Joint | 452 (12.82%) | 412 (11.68%) |
| Neck pain | 160 (4.54%) | 402 (11.40%) |
| Muscle disorders | 107 (3.03%) | 382 (10.83%) |
| No diagnoses | 616 (17.47%) | 370 (10.49%) |
| Autoimmune | 454 (12.87%) | 321 (9.10%) |
| Other osteoarthritis | 278 (7.88%) | 298 (8.45%) |
| Tendinitis (lower extremities) | 217 (6.15%) | 237 (6.72%) |
| Knee osteoarthritis | 138 (3.91%) | 225 (6.38%) |
| Fibromyalgia and unspecified tendinitis | 210 (5.95%) | 217 (6.15%) |
| Peripheral neuropathy | 119 (3.37%) | 198 (5.61%) |
| Other joint disease | 298 (8.45%) | 183 (5.19%) |

#### 3.2.2. Bivariate and multivariate analysis

After re-classifying predictors according to their prevalence, 13 disease groups remained. The list of variables assessed in the bivariate analyses can be found in Supplementary Material *Bivariate and multivariates analyses* section.

Table 3 shows the multivariate analysis results. As it can be observed, diagnoses related to highly disabling mechanical conditions (i.e., back pain, muscle disorders, tendinitis) were associated with a higher probability of occupation collection. However, mild distress and slight disability do not appear to have a significant impact on the likelihood of occupational information collection.

**Table 3.** Multivariate analysis results after conducting hybrid stepwise feature selection. Second objective: association between occupation collection and patients’ clinic and demographic characteristics.

| Variable | OR | p-value |
| --- | --- | --- |
| (Intercept) | 0.97 (0.85-1.1) | 0.642 |
| Back pain | 1.94 (1.69-2.23) | 2.30E-20 |
| Tendinitis (upper extremities) | 1.8 (1.56-2.07) | 3.40E-16 |
| Muscle disorders | 2.6 (2.05-3.32) | 9.30E-15 |
| Peripheral neuropathy | 2.06 (1.62-2.63) | 4.85E-09 |
| Osteoarthritis of knee | 1.84 (1.47-2.31) | 1.60E-07 |
| Neck pain | 1.64 (1.32-2.03) | 6.02E-06 |
| No diagnosis | 0.77 (0.66-0.9) | 0.001 |
| Other joint disease | 0.77 (0.63-0.94) | 0.011 |
| Pain in joint | 1.16 (0.99-1.36) | 0.062 |
| **Distress** |  |  |
| No distress | Ref. |  |
| Mild distress | 0.72 (0.63-0.82) | 1.02E-06 |
| Moderate and severe distress | 0.89 (0.74-1.08) | 0.254 |
| **Disability** |  |  |
| No disability | Ref. |  |
| Slight disability | 0.78 (0.7-0.88) | 7.20E-05 |
| Disability | 1 (0.85-1.18) | 0.998 |

### 3.3. Association between occupation and patient’s diagnosis

A total of 402 ESCO codes were used to manually normalise the data. The codes were later re-classified into 34 categories, see Supplementary Excel File ESCO classification and Supplementary Table 13. Figure 2 shows the OR of occupations that were significantly associated with each diagnosis, and Supplementary Table 14 present the results of the multivariate analysis on the association between the occupation and the patient’s diagnosis. As it can be appreciated, “*cleaners and helpers*” and “*social work associte professionals*” exhibit similar patterns: they are associated with mechanical diseases such as back pain, Osteoarthritis of the knee (OAk) or muscle disorders. On one hand, “*electrical and electronic trades workers*”, as well as “*sales and purchasing agents and brokers*”, are more commonly associated with OAk. On the other hand, “*sports and fitness workers*” and “*garment and related trades workers*” are predominantly associated with other joint diseases. “*Food preparation assistants*” tend to suffer from peripheral neuropathy. “*Customer service clerks*” are exclusively linked to autoimmune pathologies. Finally, “*Hairdressers, beauticians, and related workers*” occupation play a similar behaviour in autoimmune and tendinitis of the upper extremities diseases.

**Figure 2:**
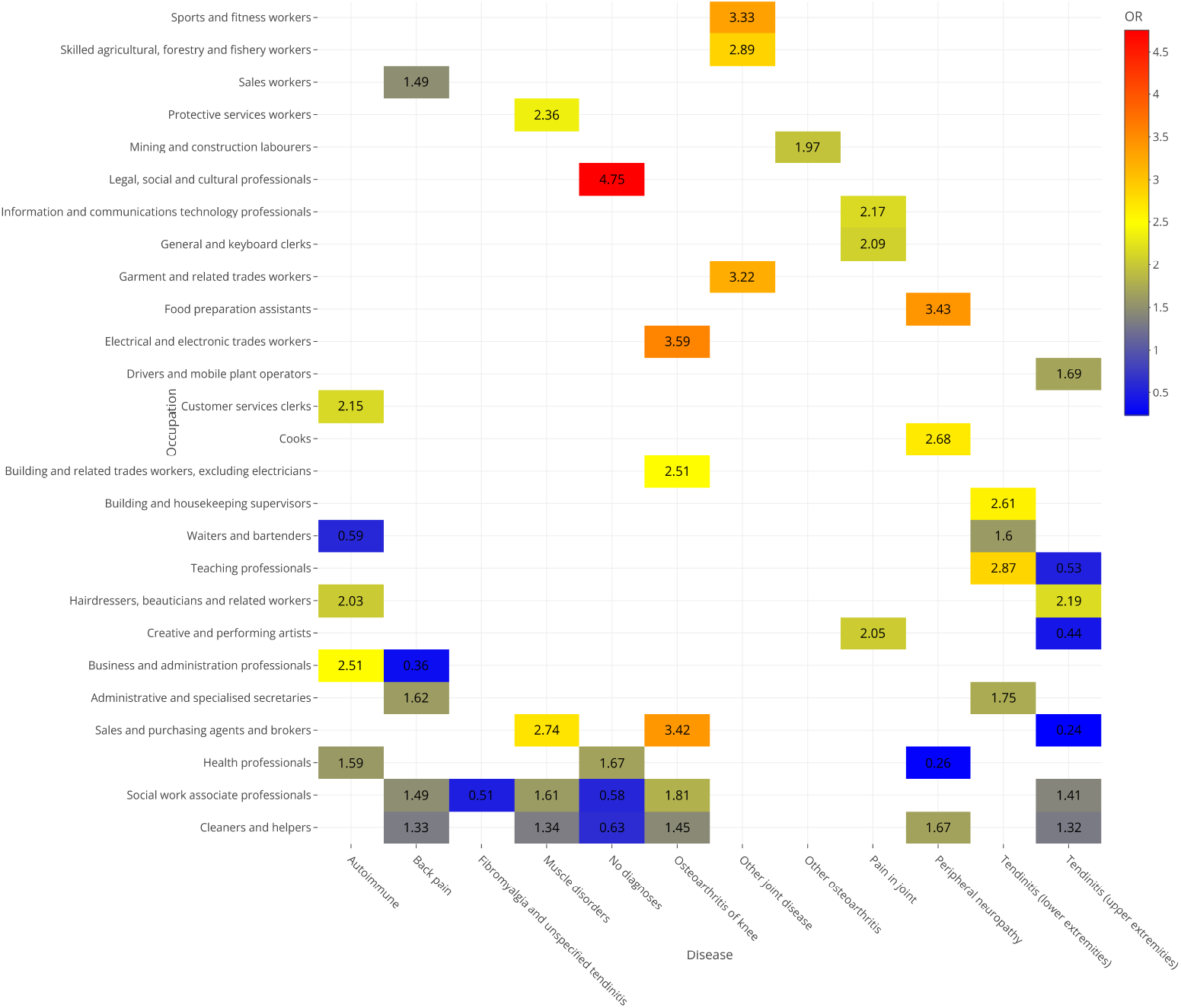
Heatmap showing the OR of occupations that were significantly associated with diagnoses. Neck pain did not contain any associated occupation and therefore was not included. The multivariate analyses results can be found in Supplementary Table 14

## 4. Discussion

In this work, we have explored the occupation collection prevalence in a departmental EHR; the sociodemographic characteristics of patients from whom occupation information was gathered compared to those without such data; and the association between occupation and patient’s diagnosis. This has been made possible thanks to the use of novelty NLP DL architectures (i.e., transformers) for NER. The use of AI techniques by the rheumatology research community has been described elsewhere [40, 41]. As reported in previous studies, this SDOH is collected in a very low proportion in the EHR. In the biomedical literature, occupation recognition has been recently studied in depth in [25], and this task has been commonly addressed simultaneously with the recognition of other SDOH [42, 43, 44, 45, 46]. A review published in 2021 [10] covered the approaches used for extracting different SDOH. Of 6,402 publications, 82 met the inclusion criteria and only 7 included occupational information. The approaches used in these seven articles consisted of rule-based algorithms. The authors in [47] presented a 10-step method for developing and validating an application to text-mining occupations from psychiatric clinical notes. A noteworthy finding was that the percentage of patients with an occupation recorded increased from 14% to 57% when considering unstructured fields. On the other hand, Dehghan et al [48] developed a large occupation dictionary used to identify occupation mentions on public and non-public clinical datasets from different institutions and countries. Researchers combined rules and ML algorithms for that purpose. However, when using rules, the results are highly dependent on their quality, have difficulties in dealing with negation, uncertainty, and ambiguity, do not scale well with increasing data size, and lack flexibility and generalizability. Moreover, there is always the risk of rules becoming outdated due to the evolving nature of biomedical language. Similarly, ML approaches also have drawbacks; they require the time-consuming creation of handcrafted features and a large amount of labeled data to perform well [49]. To overcome these drawbacks, approaches centred on DL methods have gained relevance. For instance, as part of the 2022 National NLP Clinical Challenges (n2c2) task 2, authors in [50] trained a transformer model with SHAC corpus achieving a 0.85-0.90 F1-score when extracting the employment status. In this work, our F1-score is lower, however it is noteworthy to note that corpus and other NLP resources are not easily accessible in languages other than English [51], and the training data is scarce.

As shown in objective 2, the presence of mechanical pathologies promotes the occupation collection. However, this might be influenced by the fact that mechanical disorders are those more related to work disability and sick leave. Rheumatologists from the HCSC have developed and implemented different work disability programmes over the last decades in different health areas of Madrid, which could h ave e nriched t he collection of occupation for these pathologies [52].

The relationship between cleaners and mechanical pathologies (e.g., back, neck, shoulders, elbows, hands, and lower limbs), has been previously reported by the EU-OSHA [53]. In [54, 55] authors also described the association between RMDs, such as upper limb disorders, and this profession. Moreover, the association between autoimmune diseases such as RA and hairdressers and beauticians was also described more than twenty years ago [56]. It is believed that direct contact with toxic substances may be involved. On their behalf, Ilar et at reported a moderately increased risk of ACPA+ RA in assistant nurses [57]. Readymade garment workers and joint disease relation has also been shown in the past [58]. Futhermore, distal ulnar neuropathy has been reported in chefs [59]. All these findings are aligned with our results. Eventually, there are other significant associations such as OAk and electrical workers or brokers that have not been described and that could be relevant to examine further.

### 4.1. Strenghts

One of the primary strengths of this study is its innovative and novel approach to detect occupations in rheumatology-related unstructured texts using transformers models. Moreover, to the best of our knowledge there has been no previous study attempting to demonstrate the relationship between occupations and diseases in a rheumatology outpatient electronic medical record. Therefore, this study could serve as a starting point for further exploration of occupation as a SDOH and its implications in rheumatology. Eventually, the findings of this study are consistent with existing literature on the subject.

### 4.2. Limitations

This study has certain technical limitations:

- The NER models were not fine-tuned on data from the HCSC-MSKC. By using data from this cohort, the model could have learned local expressions, syntax nuances or abbreviations that could have resulted in higher performance. The reason why fine-tuning was not performed using HCSC-MSKC data was because a cloud computing environment, not GDPR compliant, was employed for training. Nevertheless, in low-budget settings and in centres without access to specialized hardware, NER models with acceptable performance can be trained using publicly available corpus resources and cloud computing services. However, rheumatology-specific corpus for training AI systems are scarce. Recent efforts, such as RheumaLpack [60], are trying to solve this.
- The MEDDOPROF training set did not contain any rheumatology clinical notes, see Supplementary Table 1. However, as occupation entities are not exclusively collected in a concrete medical speciality, we assume that this did not significantly affect the models performance.
- In this work, two models for two different tasks were trained and their outputs combined. Other authors have experimented with the cross-concatenation of the occupation classes and the family relation classes, training a single model [61]. However, a reduction in performance of 10% was noticed.
- Automatic occupation normalization task to a common terminology was not addressed in this work. Conversely, we manually normalized each recognised entity.
- Not all rheumatologists involved in HCSC-MSKC collect occupations with the same frequency, but the distribution of new patients is random among them (i.e., they will not see a pathology more frequently with respect to the others).

## 5. Conclusion

We have accurately identified occupation mentions in real-life rheumatology clinical notes using novel NLP approaches. The acquisition of occupation-related information is only collected in a small percentage of patients, around 10%, posing significant challenges to its integration into clinical decision-making processes. Diagnoses related to highly disabling mechanical pathologies were associated with a higher probability of occupation collection by the phyisician. Ultimately, we have shown the association between occupation and more than ten diagnoses. Our findings largely align with existing literature, although we have also identified some unreported associations between occupations and RMDs.

## Supporting information

Supplementary Excel File ESCO Classification

Supplementary Excel File Predictors Classification

Supplementary Material MI-CLAIM checklist

## Data Availability

The transformers' models used in this study are openly available in Hugging Face https://huggingface.co/HCSCRheuma/Occupations

## A. Appendix

### CRediT authorship contribution statement

**Alfredo Madrid-García:** Conceptualization of this study, methodology, annotation, coding, writing (original draft preparation), feature engineering, review. **Inés Pérez-Sancristóbal:** Annotation, feature engineering. **Leticia-Leon:** Conceptualization of this study, review. **Lydia-Abásolo:** Conceptualization of this study, review. **Benjamín Fernández-Gutiérrez:** Conceptualization of this study. **Luis Rodríguez-Rodríguez:** Conceptualization of this study, methodology, feature engineering, review.

All of the authors were involved in the drafting and/or revising of the manuscript.

### Data availability statement

The transformers’ models used in this study are openly available in Hugging Face https://huggingface.co/HCSCRheuma/Occupations [30].

### Supplementary material files

- Supplementary Material: expanded methodology section with additional supplementary tables and figures.
- Supplementary Excel File Predictors Classification: classification of clinical and demographic predictors into new categories to conduct logistic regression analysis as stated in objective 2 and 3.
- Supplementary Excel File ESCO Classification: classification of occupations into new categories to conduct logistic regression analysis as stated in objective 3.
- Supplementary Material MI-CLAIM checklist

