## Supplementary Material MI-CLAIM checklist for "Occupation Recognition and Exploitation in Rheumatology Clinical Notes: Employing Deep Learning Models for Named Entity Recognition and Knowledge Discovery in Electronic Health Records"

**Checklist for supervised clinical ML study**

| **Before paper submission** | | | |
| --- | --- | --- | --- |
| **Study design (Part 1)** | **Completed:**  **page number** | | **Notes if not completed** |
| The clinical problem in which the model will be employed is clearly detailed in the paper. | ☒ | 2 | *“Until today, studies that focus on the patient's occupation are scarce, and even more, studies that attempt to identify occupation mentions in EHR using advanced DL approaches, for knowledge discovery.*  *To address this gap and with the aim of characterizing the patient's occupation, we proposed the use of transformers for occupation NER in a rheumatology departmental EHR”* |
| The research question is clearly stated. | ☒ | 2 | *“Until today, studies that focus on the patient's occupation are scarce, and even more, studies that attempt to identify occupation mentions in EHR using advanced DL approaches, for knowledge discovery.*  *To address this gap and with the aim of characterizing the patient's occupation, we proposed the use of transformers for occupation NER in a rheumatology departmental EHR"* |
| The characteristics of the cohorts (training and test sets) are detailed in the text. | ☒ | Supp material | Supplementary Material Model development section and Supplementary Tables 1-4 |
| The cohorts (training and test sets) are shown to be representative of real-world clinical settings. | ☐ |  | Not applicable. The training and validation sets are comprised of different Spanish public clinical notes and cases published in the MEDDOPROF corpus. |
| The state-of-the-art solution used as a baseline for comparison has been identified and detailed. | ☒ | 5 | Discussion section. “*For instance, as part of the 2022 n2c2 task 2, authors trained a transformer model with SHAC corpus achieving a 0.85-0.90 F1-score when extracting the employment status.”* However, we have not use this result as the baseline because different problems are addressed and direct comparisons can’t be established. |
| **Data and optimization (Parts 2, 3)** | **Completed:**  **page number** | | **Notes if not completed** |
| The origin of the data is described and the original format is detailed in the paper. | ☒ | Supp material | Supplementary material “*Hence, two different folders with notes in .txt and annotations in .ann extension are provided, one for each task (i.e., MEDDOPROF-NER and MEDDOPROF-CLASS).”* |
| Transformations of the data before it is applied to the proposed model are described. | ☒ | Supp  material | Supplementary Material Data manipulation and pre-processing section |
| The independence between training and test sets has been proven in the paper. | ☒ | Supp  material | Supplementary Material MEDDOPROF corpus description section *“The corpus is split into two subsets: training (n = 1,500) notes and validation (n = 344) notes”* On the other hand, 2,000 clinical notes from HCSC-MSKC were randomly chosen for annotation to build a gold standard, this process is described in more detail in the Supplementary Material annotation process and gold standard section. These notes comprised the test set. The split was given by MEDDOPROF authors. |
| Details on the models that were evaluated and the code developed to select the best model are provided. | ☒ | Supp  material | Supplementary Material section “Model development” and Supplementary Table 4 |
| Is the input data type structured or unstructured? | ☐ Structured ☒Unstructured | | |
| **Model performance (Part 4)** | **Completed:**  **page number** | | **Notes if not completed** |
| The primary metric selected to evaluate algorithm performance (eg: AUC, F-score, etc) including the justification for selection, has been clearly stated. | ☒ | 2 | Methodology section “The evaluation metrics used to assess the performance of the models were precision, recall and F1-score” |
| The primary metric selected to evaluate the clinical utility of the model (eg PPV, NNT, etc) including the justification for selection, has been clearly stated. | ☐ |  | Not applicable. We have shown a model for structuring clinical data. The clinical utility should be measured in final applications. |
| The performance comparison between baseline and proposed model is presented with the appropriate statistical significance. | ☐ |  | No comparison between a baseline model and the one proposed have been done. See item “The state-of-the-art solution used as a baseline for comparison has been identified and detailed.” |
| **Model Examination (Parts 5)** | **Completed:**  **page number** | | **Notes if not completed** |
| Examination Technique 1^a^ | ☐ |  | Not applicable |
| Examination Technique 2^a^ | ☐ |  | Not applicable |
| A discussion of the relevance of the examination results with respect to model/algorithm performance is presented. | ☐ |  | Not applicable |
| A discussion of the feasibility and significance of model interpretability at the case level if examination methods are uninterpretable is presented. | ☐ |  | Not applicable |
| A discussion of the reliability and robustness of the model as the underlying data distribution shifts is included. | ☐ |  | Not applicable |
| *Common examination approaches based on study type:  * For studies involving exclusively structured data coefficients and sensitivity analysis are often appropriate  * For studies involving unstructured data in the domains of image analysis or NLP: saliency maps (or equivalents) and sensitivity analysis are often appropriate |  |  | Not applicable |
| **Reproducibility (Part 6): choose appropriate tier of transparency** | | | **Notes** |
| Tier 1: complete sharing of the code | | ☐ |  |
| Tier 2: allow a third party to evaluate the code for accuracy/fairness; share the results of this evaluation | | ☐ |  |
| Tier 3: release of a virtual machine (binary) for running the code on new data without sharing its details | | ☒ | The trained models are available at <https://huggingface.co/HCSCRheuma/Occupations>  doi:10.57967/hf/0947 |
| Tier 4: no sharing | | ☐ |  |

PPV: Positive Predictive Value

NNT: Numbers Needed to Treat

^a^ Common examination approaches based on study type: for studies involving exclusively structured data, coefficients and sensitivity analysis are often appropriate; for studies involving unstructured data in the domains of image analysis or natural language processing, saliency maps (or equivalents) and sensitivity analyses are often appropriate. Select 2 from this list or chose an appropriate technique, document each technique used on the appropriate line above.
