## Supplementary Material for "Occupation Recognition and Exploitation in Rheumatology Clinical Notes: Employing Deep Learning Models for Named Entity Recognition and Knowledge Discovery in Electronic Health Records"

García et al. (2024)

The main sections of this supplementary material correspond to each of the objectives described in the main manuscript.

### 1 Performance of NER models in HCSC-MSKC gold standard set

#### 1.1 MEDDOPROF corpus description

MEDDOPROF corpus was described in depth in [1]. This corpus is comprised of clinical cases and notes from different medical specialities, see Supplementary Table 1. The corpus is split into two subsets: training (n = 1,500) notes and validation (n = 344) notes. This corpus was created in the context of a shared-task [2], and contains two set of annotations in brat rapid annotation tool (BRAT) format. The first set of notes is known as MEDDOPROF-NER and contains annotations related to:

- Professions: occupations that provide a person with an income or livelihood, including conventional professions, civil servants, public employees, new professions, and illegal professions. 'Ex' and 'Co' prefixes are considered part of the profession.
- Working status: including homemaker; retired; unemployed; unpaid caregiver; student, PhD student, apprentice, competitive examinations student; under temporary employment regulation; self-employed; on maternity/paternity leave; slave; prisoner, homeless, pauper; worker; other unspecified professional; refugee; hourly, full-time, part-time job; military service; military veteran; and co-worker or colleague.
- Activities: non-remunerated professions such as non-professional athlete/entertainer; unpaid community positions; activist; volunteer; guru or gamer.

The second set of notes is known as MEDDOPROF-CLASS and contains annotations related to:

- Patient: main actor of the clinical note.
- Family member: patient's relative
- Health professional: healthcare professional who interacts with the patient, namely, primary and secondary doctors, nurses, and assistant nurses.
- Others: mention of other persons not included in any of the above categories

Hence, two different folders with notes in .txt and annotations in .ann extension are provided in MEDDOPROF corpus, one for each task (i.e., MEDDOPROF-NER and MEDDOPROF-CLASS). Supplementary Table 2 shows the train and validation set statistics and Supplementary Table 3 shows the distribution of the annotations in the MEDDOPROF corpus.

#### 1.2 Data manipulation and pre-processing

Bidirectional encoder representations from transformers (BERT)-based models are known for their low pre-processing requirements and a decline in performance if conventional natural language processing (NLP) pre-processing techniques like stemming or stopwords removal are applied. The following steps were conducted to transform the annotated MEDDOPROF data into the expected BERT input format. These steps have been described in the literature [3]:

- .ann to BIO: *brat_to_conll.py* script from NeuroNER [4] is used to transform the annotations in standoff BRAT format to BIO format. In brief, B stands for first token in an entity, I for other tokens in an entity, and O for every token not included in an entity. These tags locates the boundaries of an entity in a sentence. The BIO tags are followed by others tags that indicates the type of entity, In this work, these tags are professions, working status, activities and/or patient, family member, health professional, others. Hence, this schema provides two kinds of tags: the position of an entity (i.e., B, I, O) in a token and the entity type.

- Input text length: to handle the length of the input text, and the maximum length of the BERT-based models, the clinical notes were split into independent sentences and the models were trained with all the information contained in the clinical note

- Text to tensor: The input data is tokenized according to the tokenizer implemented by the pre-trained language model (PLM). After tokenization, the subtokens receive the same BIO tag that the original unsplit token. Besides, as the input text can be of varying lengths, padding is done to homogenize the length of all of them. Next, attention masks are created to ignore padding labels. Finally, the data are converted to torch tensors.

##### 1.3 Model development

The model used in this work, biomedical RoBERTa-based pre-trained language model (PLM) with Spanish corpus, and its hyperparameters were set after assessing different hyperparameters and models such as BERT, AlBERT, DistilBERT or RoBERTa. This was discussed in [1]. In this work, the training set of the MEDDOPROF corpus was split into two subsets, training (80%) and validation (20%). The hyperparameters of the best performing model were identified and used to train the final models used in the current research, see Supplementary Table 4.

Python 3.8.16 was used to carry out the experiments and Google Colab was used as the cloud environment for conducting the training. The models were fine-tuned with a Nvidia Tesla T4 GPU.

##### 1.4 Annotation process and gold standard

2,000 first visit notes from the HCSC-MSKC cohort were randomly selected and annotated to build a gold standard and to assess the model's performance before making inference, locally, on the rest of the notes, Supplementary Table 5. These 2,000 notes were annotated by two annotators, AMG and IPS, using brat rapid annotation tool (BRAT) and following MEDDOPROF corpus annotation guidelines [5]. The inter-annotator agreement (IAA) between the two annotators was measured using the bratiaa python package accessible through GitHub [6]. The characteristics of the gold standard set are summarized in Supplementary Table 6. The distribution of the entities among the different labels can be seen Supplementary Table 7.

Initially, the token and instance IAA mean F1 was 0.668 and 0.687 respectively. After computing this score with bratiaa, both annotators met to resolve discrepancies and correct errors (e.g., not detected entities and/or annotation errors) with the aim of building a robust gold standard. Once the discrepancies were sorted, the entities distribution, Supplementary Table 7, and the confusion matrices for both tasks, Supplementary Table 8 and 9, as well as the combined confusion matrix, Supplementary Table 10, were studied. The results of the evaluation library, seqeval, are shown in Supplementary Table 11. As it can be appreciated, the patient identification is the limiting task of the combined model, this is, the hardest task of the two proposed.

As shown in Supplementary Table 10, six and eight entities recognised by the model as "Profession" and as "Patient" were actual patient occupations (8.4%). We therefore opted to study these entities as well.

### 2 Demographic and clinical characteristics that influence occupation collection

#### 2.1 Predictions in the HCSC-MSKC notes

Supplementary Table 12 shows the number of entities recognised by the models, 33,292, when using all the notes from HCSC-MSKC before accounting for selection bias. Of them, 7,314 belongs to "Patient" (n = 2,307), "Profession" (n = 1,305) or "Profession-patient" (n = 3,702). After manual review, 189 (8.19%) and 45 (3.45%) notes with only "Patient" or "Profession" entities were actually "Profession-patient" and therefore, recovered.

#### 2.2 Matching visits

Each visit with an occupation mention is paired with an available visit from a patient with no occupation mentions that has the closest propensity score to it. From this point, there were two options:

1. Visits without occupation mentions can come from the same controls (i.e., number of visits without occupation mention is greater than the number of patients without occupation mentions)

2. Each control can only provide one visit (i.e., number of visits without occupation mention is the same as the number of patients without occupation mentions)

Both options were considered and analyses were repeated for both scenarios. Finally, the first approach was chosen as propensity scores are more similar between the comparison groups.

Eventually, Supplementary Figure 4 shows the love plot after balancing. Balance was achieved for all the covariates with a standardized mean difference $< 0.1$.

#### 2.3 Bivariate and multivariates analyses

The variables included in the bivariate and multivariate analyses were related to quality of life measures (n = 3; distress, disability, Rosser) and diagnoses (n = 13; back pain, tendinitis (upper extremities), pain in joint, neck pain, muscle disorders, no diagnosis, autoimmune, other osteoarthritis, tendinitis (lower extremities), osteoarthritis of knee, fibromyalgia and unspecified tendinitis, peripheral neuropathy and other joint disease).

After bivariate analyses, rosser, fibromyalgia and unspecified tendinitis, other osteoarthritis, and tendinitis (lower extremities) variables were excluded from subsequent analysis as their p-value was $> 0.15$.

After multivariate analyses following an hybrid stepwise approaches optimising the AIC, autoimmune disease was excluded.

### 3 Association between occupation and patient's diagnosis

Supplementary Table 13 shows the prevalence of the 34 diagnosis categories. The number of occupations categories found in the 3,527 visits was 3,636.

Supplementary Table 14 shows the result of the multivariate analysis for assessing the association between occupation and patient's diagnosis.

### Supplementary Tables

#### MEDDOPROF corpus related tables

Supplementary Table 1: MEDDOPROF clinical notes specialities. Other I: includes all clinical cases starting with SXXXX-. Other II: includes all clinical cases starting with XXXXXXXX_ES

| **Speciality**<br>**N (%)** | **total**<br>**n = 1,844** | **train**<br>**n = 1,500 (0.81)** | **test**<br>**n = 344 (0.19)** |
|---|---|---|---|
| Psychiatry | 560 | 484 (0.86) | 76 (0.14) |
| Labour | 233 | 81 (0.35) | 152 (0.65) |
| Internal medicine | 229 | 207 (0.9) | 22 (0.1) |
| Oncology | 194 | 175 (0.9) | 19 (0.1) |
| Primary care | 93 | 86 (0.92) | 7 (0.08) |
| Dermatology | 87 | 77 (0.89) | 10 (0.11) |
| Infectology | 65 | 58 (0.89) | 7 (0.11) |
| Neurology | 63 | 54 (0.86) | 9 (0.14) |
| Other II | 58 | 50 (0.86) | 8 (0.14) |
| Emergency | 35 | 34 (0.97) | 1 (0.03) |
| Radiology | 31 | 27 (0.87) | 4 (0.13) |
| Otorhinolaryngology | 28 | 26 (0.93) | 2 (0.07) |
| Allergology | 25 | 24 (0.96) | 1 (0.04) |
| Odontology | 24 | 22 (0.92) | 2 (0.08) |
| Ophthalmology | 24 | 22 (0.92) | 2 (0.08) |
| COVID | 20 | 19 (0.95) | 1 (0.05) |
| Urology | 20 | 16 (0.8) | 4 (0.2) |
| Other I | 19 | 16 (0.84) | 3 (0.16) |
| Tropical medicine | 18 | 15 (0.83) | 3 (0.17) |
| Endocrinology | 10 | 7 (0.7) | 3 (0.3) |
| Rheumatology | 8 | 0 (0) | 8 (1) |

Supplementary Table 2: Number or documents, annotations, unique codes, and sentences in the MEDDOPROF corpus. Table extracted from IberLEF 2021 - MEDDOPROF video

| | Documents | Annotations | Sentences | Tokens |
|---|---|---|---|---|
| Train | 1,500 | 3,658 | 49,114 | 1,075,655 |
| Validation | 344 | 1,085 | 9,513 | 215,531 |
| Total | 1,844 | 4,743 | 58,627 | 1,291,186 |

Supplementary Table 3: Proportion of entities in the MEDDOPROF corpus. In parentheses, train and test proportions. Table extracted from [1]

| | Patient | Family | Health Prof. | Other | Total |
|---|---|---|---|---|---|
| Profession | 1,158<br>(876-282) | 134<br>(105-29) | 1,525<br>(1,231-294) | 410<br>(316-94) | 3,227 (68.04%)<br>(2,528-699) |
| Empl. Status | 1,047<br>(754-293) | 119<br>(97-22) | 0 | 203<br>(160-43) | 1,369 (28.86%)<br>(1,011-358) |
| Activity | 122<br>(105-17) | 7<br>(5-2) | 0 | 18<br>(9-9) | 147 (3.10%)<br>(119-28) |
| Total | 2,327 (49.06%)<br>(1,735-592) | 260 (5.5%)<br>(207-53) | 1,525 (32.14%) | 631 (13.29%)<br>(485-146) | 4,743<br>(3,658-1,085) |

Supplementary Table 4: Models' parameters

| **PLM Model** | **Learning rate** | **Batch size** | **Epochs** | **Max token length** | **Optimizer** | **Max clip grad norm** | **Epsilon** |
|---|---|---|---|---|---|---|---|
| RoBERTa base biomedical clinical es | 2e-05 | 8 | 10 | 510 | AdamW | 1 | 1e-08 |

#### HCSC-MSKC related tables

Supplementary Table 5: Number of randomly selected clinical notes per year composing gold standard set (HCSC-MSKC)

| Year | Number of notes |
|---|---|
| **2007** | 348 |
| **2008** | 381 |
| **2009** | 228 |
| **2010** | 100 |
| **2011** | 101 |
| **2012** | 100 |
| **2013** | 115 |
| **2014** | 86 |
| **2015** | 127 |
| **2016** | 101 |
| **2017** | 313 |

Supplementary Table 6: Number of documents, annotations, and sentences in the gold standard set (HCSC-MSKC)

| Corpus | Documents | Annotations | Sentences | Tokens |
|---|---|---|---|---|
| Gold standard set (HCSC-MSKC) | 2,000 | 898 | 15,306 | 202,173 |

Supplementary Table 7: Proportion of entities in the gold standard set (HCSC-MSKC)

| | Patient | Family | Health Prof. | Other | Total |
|---|---|---|---|---|---|
| Profession | 167 | 5 | 579 | 1 | 752 (83.74%) |
| Empl.Status | 103 | 1 | 0 | 0 | 104 (11.58%) |
| Activity | 42 | 0 | 0 | 0 | 42 (4.68%) |
| Total | 312 (34.74%) | 6 (<1%) | 579 (64.48%) | 1 (<1%) | 898 |

Supplementary Table 8: Occupation recognition task confusion matrix. Gold standard set (HCSC-MSKC). BIO schema. ACT: Activity, PRO: Profession, WS: Working status

| | | **Actual (Gold Standard)** | | | | | | | |
|---|---|---|---|---|---|---|---|---|---|
| | | B-ACT | B-PRO | B-WS | I-ACT | I-PRO | I-WS | O | **support** |
| **Predicted** | B-ACT | 13 | 2 | 0 | 10 | 0 | 0 | 17 | **42** |
| | B-PRO | 0 | 595 | 1 | 0 | 5 | 0 | 151 | **752** |
| | B-WS | 0 | 7 | 48 | 0 | 2 | 2 | 45 | **104** |
| | I-ACT | 4 | 0 | 0 | 29 | 3 | 0 | 43 | **79** |
| | I-PRO | 0 | 8 | 0 | 0 | 332 | 2 | 50 | **392** |
| | I-WS | 0 | 0 | 5 | 3 | 19 | 61 | 81 | **169** |
| | O | 31 | 34 | 9 | 41 | 36 | 22 | 200462 | **200635** |
| | total predicted | **48** | **646** | **63** | **83** | **397** | **87** | **200849** | **202173** |

Supplementary Table 9: Identification of the actor to which the occupation belongs confusion matrix. Gold standard set (HCSC-MSKC). BIO schema. ACT: Activity, FAM: Family member, HEA: Health professional, OTH: Other, PAT: Patient, PRO: Profession, WS: Working status

| | | **Actual (Gold standard)** | | | | | | | | | |
|---|---|---|---|---|---|---|---|---|---|---|---|
| | | B-FAM | B-OTH | B-PAT | B-HEA | I-FAM | I-OTH | I-PAT | I-HEA | O | support |
| **Predicted** | B-FAM | 2 | 0 | 0 | 4 | 0 | 0 | 0 | 0 | 0 | **6** |
| | B-OTH | 0 | 1 | 0 | 0 | 0 | 0 | 0 | 0 | 0 | **1** |
| | B-PAT | 0 | 0 | 202 | 3 | 0 | 0 | 22 | 0 | 85 | **312** |
| | B-HEA | 0 | 2 | 1 | 469 | 0 | 0 | 0 | 1 | 106 | **579** |
| | I-FAM | 0 | 0 | 0 | 0 | 3 | 0 | 0 | 0 | 0 | **3** |
| | I-OTH | 0 | 0 | 0 | 0 | 0 | 0 | 0 | 0 | 0 | **0** |
| | I-PAT | 0 | 0 | 17 | 0 | 0 | 0 | 373 | 4 | 160 | **554** |
| | I-HEA | 0 | 0 | 0 | 1 | 0 | 4 | 0 | 72 | 6 | **83** |
| | O | 0 | 6 | 54 | 30 | 0 | 0 | 118 | 3 | 200424 | **200635** |
| | total predicted | **2** | **9** | **274** | **507** | **3** | **4** | **513** | **80** | **200781** | **202173** |

Supplementary Table 10: Occupation recognition and actor to which the occupation belongs combined confusion matrix. Gold standard set (HCSC-MSKC). ACT: Activity, FAM: Family member, OTH: Other, PAT: Patient, HEA: Health professional. WS: Working status

| | | **Actual (Gold standard)** | | | | | | | | |
|---|---|---|---|---|---|---|---|---|---|---|
| | | PROF-PAT | PROF-HEA | PROF-FAM | PROF-OTHER | WS-PAT | WS-FAM | ACT-PAT | O | support |
| **Predicted** | PROF-PAT | 115 | 1 | 0 | 0 | 5 | 0 | 1 | 20 | **142** |
| | PROF-HEA | 2 | 428 | 2 | 0 | 0 | 0 | 0 | 34 | **466** |
| | PROF-FAM | 0 | 0 | 1 | 0 | 0 | 0 | 0 | 0 | **1** |
| | PROF-OTHER | 0 | 2 | 0 | 0 | 0 | 0 | 0 | 4 | **6** |
| | WS-PAT | 0 | 0 | 0 | 0 | 35 | 0 | 0 | 14 | **49** |
| | WS-FAM | 0 | 0 | 0 | 0 | 0 | 1 | 0 | 0 | **1** |
| | ACT-PAT | 0 | 0 | 0 | 0 | 0 | 0 | 7 | 19 | **26** |
| | O | 36 | 127 | 1 | 0 | 51 | 0 | 26 | 0 | **241** |
| | PROF | 6 | 0 | 0 | 0 | 1 | 0 | 0 | 30 | **37** |
| | WS | 0 | 0 | 0 | 0 | 8 | 0 | 0 | 15 | **23** |
| | ACT | 0 | 0 | 0 | 0 | 0 | 0 | 4 | 23 | **27** |
| | PAT | 8 | 0 | 0 | 0 | 3 | 0 | 4 | 56 | **71** |
| | HEA | 0 | 21 | 1 | 0 | 0 | 0 | 0 | 18 | **40** |
| | OTHER | 0 | 0 | 0 | 1 | 0 | 0 | 0 | 2 | **3** |
| | total predicted | **167** | **579** | **5** | **1** | **103** | **1** | **42** | **235** | **1133** |

Supplementary Table 11: Precision, recall and F1 values per entity using seqeval library. Gold standard set (HCSC-MSKC)

| Task | Entity | Precision | Recall | F1 | Support |
|---|---|---|---|---|---|
| Occupation recognition | Activity | 0.21 | 0.26 | 0.23 | 42 |
| | **Profession** | 0.89 | 0.77 | 0.83 | 752 |
| | Working status | 0.62 | 0.43 | 0.51 | 104 |
| To whom the occupation belongs | Family member | 1 | 0.33 | 0.50 | 6 |
| | Other | 0.11 | 1 | 0.20 | 1 |
| | **Patient** | 0.64 | 0.59 | 0.61 | 312 |
| | Health professional | 0.92 | 0.80 | 0.86 | 579 |

Supplementary Table 12: Number of recognised entities in the whole HCSC-MSKC dataset, n = 33,292. Number of profession and/or patient related mentions, n = 7,314 belonging to 5,917 visits

| **Entity** | **n** |
|---|---|
| PROFESSION-PATIENT | 3,702 |
| PROFESSION-HEALTH PROFESSIONAL | 18,223 |
| PROFESSION-FAMILY MEMBER | 58 |
| PROFESSION-OTHERS | 309 |
| WORKING SITUATION-PATIENT | 1,588 |
| WORKING SITUATION-HEALTH PROFESSIONAL | 2 |
| WORKING SITUATION-FAMILY MEMBER | 23 |
| WORKING SITUATION-OTHERS | 39 |
| ACTIVITY-PATIENT | 1,121 |
| ACTIVITY-HEALTH PROFESSIONAL | 2 |
| ACTIVITY-FAMILY MEMBER | 3 |
| ACTIVITY-OTHERS | 3 |
| PROFESSION | 1,305 |
| WORKING SITUATION | 722 |
| ACTIVITY | 1,060 |
| PATIENT | 2,307 |
| HEALTH PROFESSIONAL | 2,616 |
| FAMILY MEMBER | 57 |
| OTHERS | 152 |

Supplementary Table 13: Prevalence of the 34 occupation categories

| **Occupation category** | **n** |
|---|---|
| Cleaners and helpers | 711 |
| Waiters and bartenders | 290 |
| Health professionals | 281 |
| Social work associate professionals | 275 |
| Sales workers | 185 |
| Administrative and specialised secretaries | 175 |
| Cooks | 162 |
| Mining and construction labourers | 122 |
| Teaching professionals | 112 |
| Building and related trades workers, excluding electricians | 104 |
| Hairdressers, beauticians and related workers | 99 |
| Drivers and mobile plant operators | 82 |
| Creative and performing artists | 79 |
| Protective services workers | 77 |
| Transport and storage labourers | 75 |
| Metal, machinery and related trades workers | 71 |
| General and keyboard clerks | 65 |
| Food processing and related trades workers | 62 |
| Business and administration professionals | 55 |
| Customer services clerks | 53 |
| Building and housekeeping supervisors | 52 |
| Science and engineering professionals | 51 |
| Sports and fitness workers | 43 |
| Information and communications technology professionals | 40 |
| Mail carriers and sorting clerks | 40 |
| Food preparation assistants | 37 |
| Sales and purchasing agents and brokers | 35 |
| Skilled agricultural, forestry and fishery workers | 35 |
| Garment and related trades workers | 31 |
| Stationary plant and machine operators | 31 |
| Legal, social and cultural professionals | 30 |
| Electrical and electronic trades workers | 29 |
| Science and engineering associate professionals | 25 |
| Child care workers and teachers' aides | 22 |

Supplementary Table 14: Multivariate analysis results after conducting hybrid stepwise feature selection. Third objective: association between occupation and patient's diagnosis

| Variable | OR | p-value |
|---|---|---|
| **Back pain** | | |
| (Intercept) | 0.57 (0.39-0.83) | 0.004 |
| Age | 0.99 (0.98-0.99) | 4.33E-04 |
| Administrative and specialised secretaries | 1.62 (1.14-2.27) | 0.006 |
| Social work associate professionals | 1.49 (1.1-1.99) | 0.009 |
| Cleaners and helpers | 1.33 (1.08-1.65) | 0.009 |
| Sales workers | 1.49 (1.06-2.08) | 0.019 |
| Business and administration professionals | 0.36 (0.13-0.83) | 0.032 |
| Sports and fitness workers | 0.41 (0.14-0.97) | 0.066 |
| Transport and storage labourers | 1.53 (0.92-2.48) | 0.093 |
| Food preparation assistants | 0.42 (0.12-1.06) | 0.103 |
| Legal, social and cultural professionals | 0.38 (0.09-1.09) | 0.116 |
| Creative and performing artists | 0.65 (0.34-1.15) | 0.162 |
| Sex (female) | 0.89 (0.74-1.08) | 0.241 |
| **Tendinitis (upper extremities)** | | |
| (Intercept) | 0.06 (0.04-0.09) | 1.41E-34 |
| Age | 1.03 (1.02-1.04) | 6.41E-13 |
| Hairdressers, beauticians and related workers | 2.19 (1.36-3.45) | 9.48E-04 |
| Sex (female) | 0.78 (0.63-0.95) | 0.014 |
| Cleaners and helpers | 1.32 (1.05-1.65) | 0.015 |
| Social work associate professionals | 1.41 (1.03-1.91) | 0.030 |
| Drivers and mobile plant operators | 1.69 (1.02-2.73) | 0.035 |
| Creative and performing artists | 0.44 (0.18-0.91) | 0.043 |
| Teaching professionals | 0.53 (0.27-0.95) | 0.045 |
| Sales and purchasing agents and brokers | 0.24 (0.04-0.79) | 0.049 |
| Legal, social and cultural professionals | 0.15 (0.01-0.69) | 0.059 |
| Electrical and electronic trades workers | 1.88 (0.83-4.04) | 0.113 |
| Science and engineering associate professionals | 0.34 (0.05-1.17) | 0.146 |
| Business and administration professionals | 0.56 (0.23-1.17) | 0.156 |
| **Muscle disorders** | | |
| (Intercept) | 0.16 (0.09-0.26) | 3.91E-12 |
| Sex (female) | 1.73 (1.31-2.29) | 1.15E-04 |
| Age | 0.98 (0.97-0.99) | 0.003 |
| Protective services workers | 2.36 (1.21-4.28) | 0.007 |
| Social work associate professionals | 1.61 (1.11-2.3) | 0.010 |
| Sales and purchasing agents and brokers | 2.74 (1.08-6.07) | 0.020 |
| Cleaners and helpers | 1.34 (1.02-1.75) | 0.037 |
| Science and engineering professionals | 0.18 (0.01-0.81) | 0.087 |
| Health professionals | 0.69 (0.42-1.09) | 0.129 |
| **Autoimmune** | | |
| (Intercept) | 0.18 (0.1-0.31) | 9.92E-10 |
| Age | 0.98 (0.97-0.99) | 0.003 |
| Hairdressers, beauticians and related workers | 2.03 (1.15-3.39) | 0.010 |
| Business and administration professionals | 2.51 (1.18-4.86) | 0.010 |
| Health professionals | 1.59 (1.08-2.28) | 0.016 |
| Customer services clerks | 2.15 (0.97-4.27) | 0.040 |

| | | |
|---|---|---|
| Waiters and bartenders | 0.59 (0.34-0.96) | 0.048 |
| Protective services workers | 0.31 (0.05-0.99) | 0.101 |
| Sex (female) | 1.22 (0.95-1.59) | 0.128 |
| **Peripheral neuropathy** | | |
| (Intercept) | 0.04 (0.02-0.09) | 3.40E-17 |
| Cooks | 2.68 (1.57-4.39) | 1.64E-04 |
| Sex (female) | 1.82 (1.26-2.67) | 0.002 |
| Cleaners and helpers | 1.67 (1.17-2.36) | 0.004 |
| Food preparation assistants | 3.43 (1.26-7.92) | 0.007 |
| Health professionals | 0.26 (0.08-0.62) | 0.008 |
| Administrative and specialised secretaries | 0.52 (0.18-1.18) | 0.164 |
| Age | 0.99 (0.98-1.01) | 0.448 |
| **Osteoarthritis of knee** | | |
| (Intercept) | 0.01 (0-0.01) | 1.07E-34 |
| Age | 1.04 (1.03-1.06) | 1.52E-07 |
| Social work associate professionals | 1.81 (1.15-2.79) | 0.008 |
| Building and related trades workers, excluding electricians | 2.51 (1.16-4.96) | 0.013 |
| Sales and purchasing agents and brokers | 3.42 (1.14-8.39) | 0.014 |
| Electrical and electronic trades workers | 3.59 (1.02-9.76) | 0.023 |
| Sex (female) | 1.49 (1.04-2.15) | 0.032 |
| Cleaners and helpers | 1.45 (1.02-2.04) | 0.034 |
| Creative and performing artists | 2.13 (0.87-4.5) | 0.067 |
| Waiters and bartenders | 0.58 (0.26-1.13) | 0.145 |
| **Neck pain** | | |
| (Intercept) | 0.08 (0.05-0.13) | 2.78E-20 |
| Sex (female) | 2.62 (1.97-3.52) | 7.11E-11 |
| Social work associate professionals | 1.41 (0.99-1.97) | 0.053 |
| Science and engineering professionals | 0.17 (0.01-0.77) | 0.078 |
| Hairdressers, beauticians and related workers | 0.5 (0.21-1.03) | 0.087 |
| Food preparation assistants | 0.2 (0.01-0.92) | 0.110 |
| Cleaners and helpers | 1.21 (0.94-1.56) | 0.138 |
| Age | 0.99 (0.98-1) | 0.276 |
| **No diagnoses** | | |
| (Intercept) | 0.37 (0.22-0.61) | 1.15E-04 |
| Age | 0.98 (0.97-0.99) | 4.84E-06 |
| Legal, social and cultural professionals | 4.75 (2.15-10.02) | 5.96E-05 |
| Health professionals | 1.67 (1.15-2.38) | 0.006 |
| Cleaners and helpers | 0.63 (0.44-0.89) | 0.010 |
| Social work associate professionals | 0.58 (0.33-0.96) | 0.047 |
| Science and engineering professionals | 1.8 (0.84-3.52) | 0.104 |
| Sports and fitness workers | 1.79 (0.79-3.64) | 0.131 |
| Hairdressers, beauticians and related workers | 1.53 (0.85-2.59) | 0.132 |
| Sex (female) | 0.93 (0.73-1.2) | 0.586 |
| **Other joint disease** | | |
| (Intercept) | 0.02 (0.01-0.05) | 2.99E-20 |
| Sex (female) | 0.32 (0.23-0.45) | 1.04E-10 |
| Age | 1.03 (1.01-1.05) | 2.77E-04 |
| Sports and fitness workers | 3.33 (1.21-7.86) | 0.010 |
| Skilled agricultural, forestry and fishery workers | 2.89 (1.13-6.51) | 0.016 |
| Garment and related trades workers | 3.22 (0.92-8.7) | 0.037 |

| | | |
|---|---|---|
| Sales and purchasing agents and brokers | 2.48 (0.82-6.11) | 0.070 |
| Cooks | 1.74 (0.89-3.16) | 0.084 |
| Teaching professionals | 0.18 (0.01-0.84) | 0.093 |
| Information and communications technology professionals | 2.27 (0.75-5.57) | 0.103 |
| Health professionals | 1.56 (0.87-2.64) | 0.114 |
| Food processing and related trades workers | 0.23 (0.01-1.08) | 0.152 |
| Cleaners and helpers | 0.66 (0.36-1.14) | 0.156 |
| **Fibromyalgia and unspecified tendinitis** | | |
| (Intercept) | 0.07 (0.04-0.13) | 5.57E-15 |
| Sex (female) | 1.4 (1.03-1.94) | 0.036 |
| Social work associate professionals | 0.51 (0.25-0.93) | 0.042 |
| Building and housekeeping supervisors | 2.23 (0.84-4.98) | 0.073 |
| Building and related trades workers, excluding electricians | 0.36 (0.06-1.17) | 0.159 |
| Age | 0.99 (0.98-1.01) | 0.431 |
| **Other osteoarthritis** | | |
| (Intercept) | 0 (0-0) | 3.84E-63 |
| Age | 1.09 (1.08-1.11) | 3.96E-30 |
| Sex (female) | 1.86 (1.39-2.52) | 4.15E-05 |
| Mining and Construction Labourers | 1.97 (1-3.65) | 0.038 |
| Business and administration professionals | 0.32 (0.05-1.07) | 0.124 |
| **Pain in joint** | | |
| (Intercept) | 0.24 (0.15-0.39) | 1.82E-08 |
| Age | 0.98 (0.97-0.99) | 8.14E-04 |
| Creative and performing artists | 2.05 (1.15-3.49) | 0.011 |
| General and keyboard clerks | 2.09 (1.1-3.72) | 0.017 |
| Information and communications technology professionals | 2.17 (0.96-4.46) | 0.046 |
| Building and housekeeping supervisors | 1.98 (0.89-3.96) | 0.068 |
| Sex (female) | 1.2 (0.96-1.51) | 0.117 |
| Protective services workers | 0.47 (0.14-1.14) | 0.142 |
| **Tendinitis (lower extremities)** | | |
| (Intercept) | 0.01 (0-0.02) | 3.68E-34 |
| Age | 1.03 (1.02-1.05) | 8.43E-06 |
| Teaching professionals | 2.87 (1.61-4.82) | 1.47E-04 |
| Sex (female) | 1.69 (1.24-2.33) | 0.001 |
| Building and housekeeping supervisors | 2.61 (1.05-5.62) | 0.023 |
| Administrative and specialised secretaries | 1.75 (1.01-2.86) | 0.033 |
| Waiters and bartenders | 1.6 (0.99-2.48) | 0.046 |
| Science and engineering associate professionals | 3.06 (0.71-9.2) | 0.077 |

### Supplementary Figures

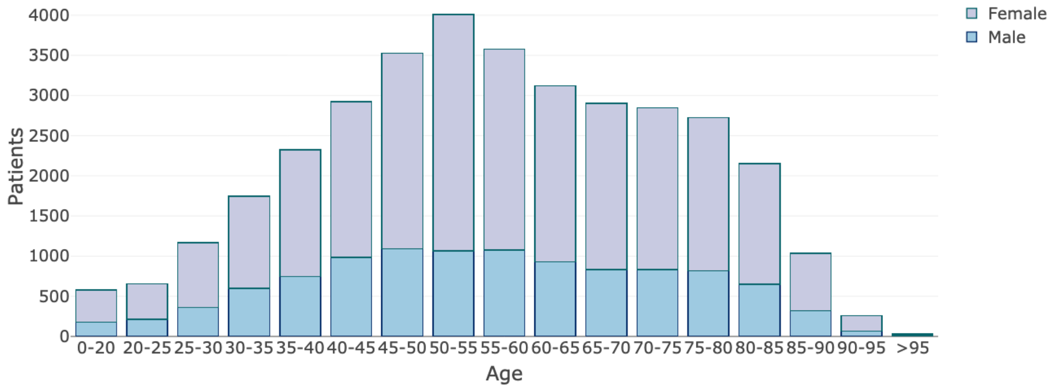


Supplementary Figure 1: HCSC-MSKC cohort age-sex distribution in first visit. The average retirement age of the Spanish population in 2017 was 65 years

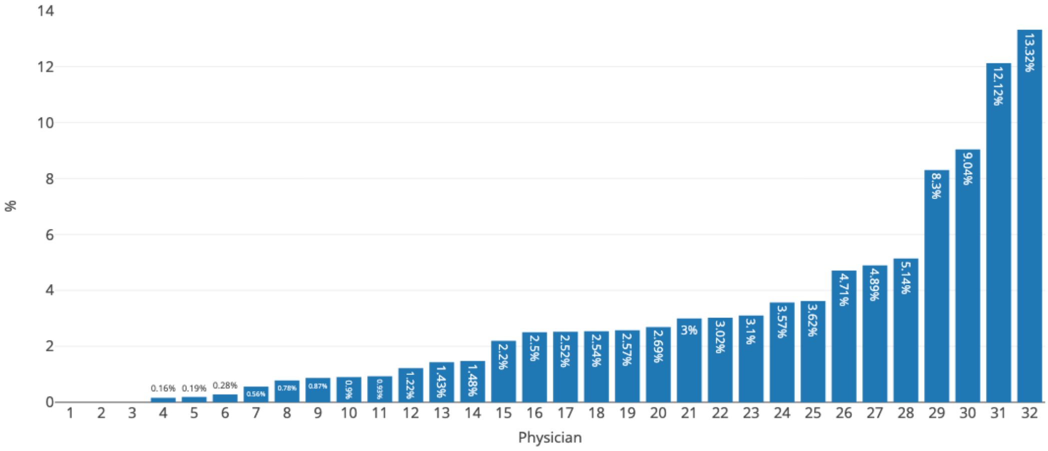


Supplementary Figure 2: Percentage of occupancy collection per physician. 117,068 visits from 35,470 patients. Of them, 3,978 visits have at least one occupation mention (from 3,723 patient)

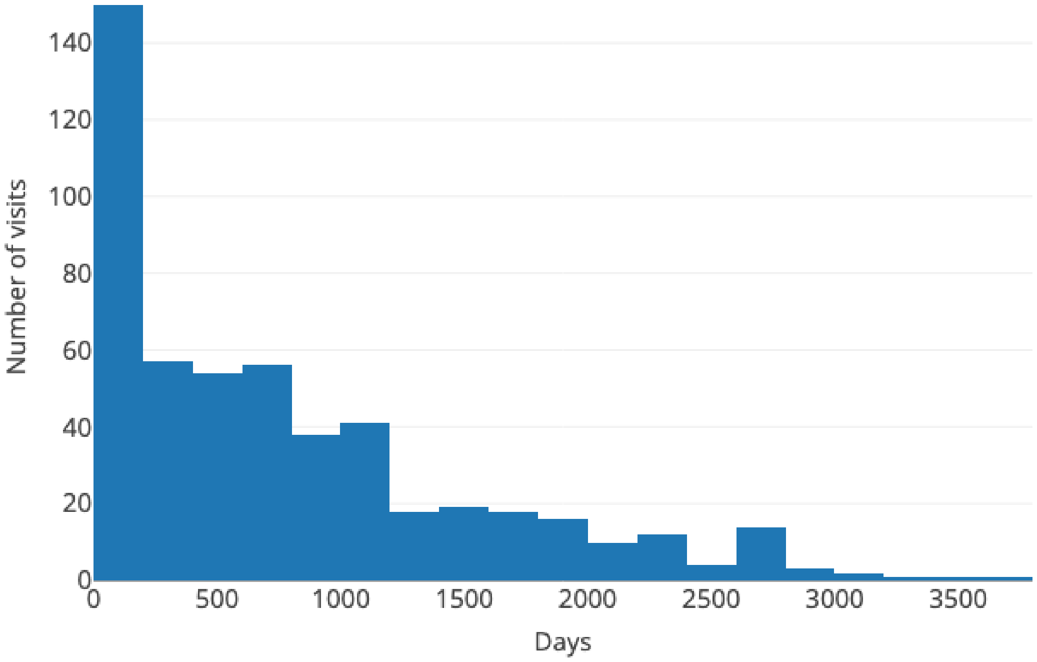


Supplementary Figure 3: Time from first patient visit without occupation to the most inmediate visit with occupation (i.e., first visit with registered occupation). n = 515 patients

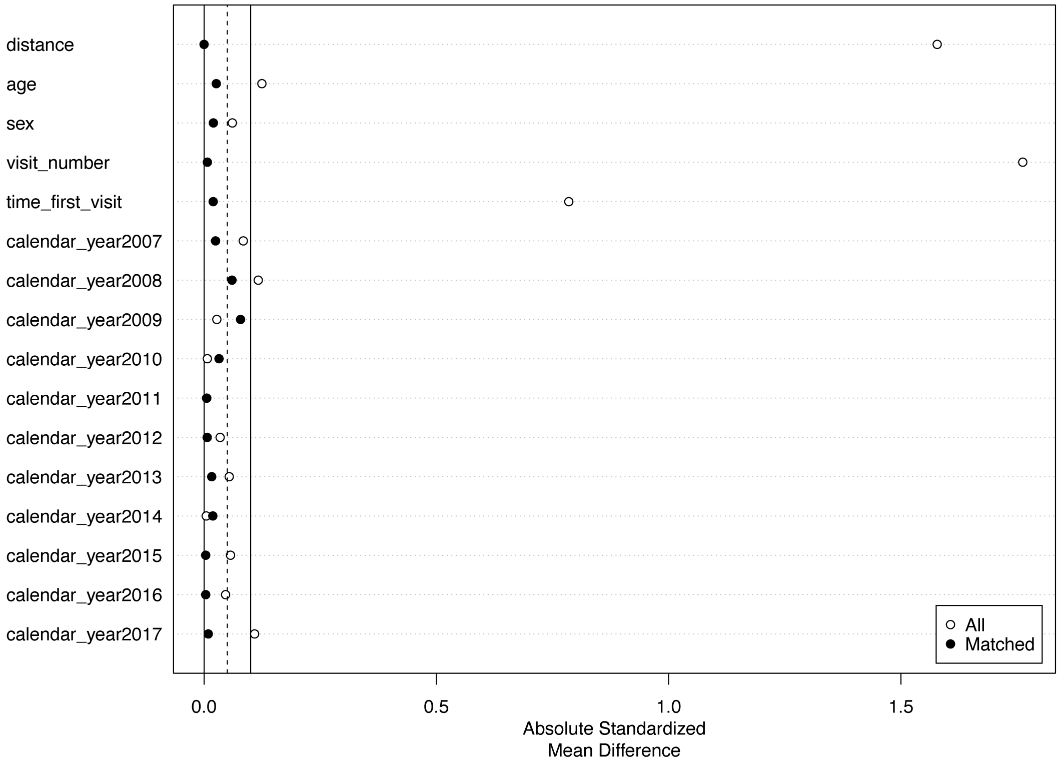


Supplementary Figure 4: Love plot for matching balance assessment

#### References


[1] Alfredo Madrid-García. "Recognition of professions in medical documentation". MA thesis. Universidad Nacional de Educación a Distancia (España), 2023.

[2] Salvador Lima-López, Eulàlia Farré-Maduell, Antonio Miranda-Escalada, et al. *NLP applied to occupational health: MEDDOPROF shared task at IberLEF 2021 on automatic recognition, classification and normalization of professions and occupations from medical texts*. 2021-09.

[3] Denis Rothman. *Transformers for Natural Language Processing: Build innovative deep neural network architectures for NLP with Python, PyTorch, TensorFlow, BERT, RoBERTa, and more*. Packt Publishing Ltd, 2021.

[4] Franck Dernoncourt, Ji Young Lee, and Peter Szolovits. "NeuroNER: an easy-to-use program for named-entity recognition based on neural networks". In: *Conference on Empirical Methods on Natural Language Processing (EMNLP)* (2017).

[5] Eulàlia Farré-Maduell, Salvador Lima-López, Antonio Miranda-Escalada, et al. *MEDDOPROF guidelines*. Apr. 2021. DOI: 10.5281/zenodo.4720833. URL: https://doi.org/10.5281/zenodo.4720833.

[6] Tobias Kolditz. *Bratiaa*. https://github.com/kldtz/bratiaa. 2019.
